# Network-based Modeling of COVID-19 Dynamics: Early Pandemic Spread in India

**DOI:** 10.1101/2021.03.16.21253772

**Authors:** Rupam Bhattachayya, Sayantan Banerjee, Shariq Mohammed, Veera Baladandayuthapani

## Abstract

Modeling the dynamics of COVID-19 pandemic spread is a challenging and relevant problem. Established models for the epidemic spread such as compartmental epidemiological models e.g. Susceptible-Infected-Recovered (SIR) models and its variants, have been discussed extensively in the literature and utilized to forecast the growth of the pandemic across different hot-spots in the world. The standard formulations of SIR models rely upon summary-level data, which may not be able to fully capture the complete dynamics of the pandemic growth. Since the disease spreads from carriers to susceptible individuals via some form of contact, it inherently relies upon a network of individuals for its growth, with edges established via direct interaction, such as shared physical proximity. Using individual-level COVID-19 data from the early days (January 30 to April 15, 2020) of the pandemic in India, and under a network-based SIR model framework, we performed state-specific forecasting under multiple scenarios characterized by the basic reproduction number of COVID-19 across 34 Indian states and union territories. We validated our short-term projections using observed case counts and the long-term projections using national sero-survey findings. Based on healthcare availability data, we also performed projections to assess the burdens on the infrastructure along the spectrum of the pandemic growth. We have developed an interactive dashboard summarizing our results. Our predictions successfully identified the initial hot-spots of India such as Maharashtra and Delhi, and those that emerged later, such as Madhya Pradesh and Kerala. These models have the potential to inform appropriate policies for isolation and mitigation strategies to contain the pandemic, through a phased approach by appropriate resource prioritization and allocation.

## 1 Introduction

Since the first reported case on November 17, 2019, in Hubei, China, the severe acute respiratory syndrome Coronavirus 2 (*SARS-CoV-2*), commonly referred to as Coronavirus Disease 2019 (COVID-19), has spread to more than 209 countries and territories. COVID-19 has afflicted more than 105 million people — resulting in over 2.6 million deaths [as of March 7, 2021, end of day], sending billions into lockdown and ravaging economies and healthcare infrastructure with each passing day. This is a pandemic at a scale that the living generation has rarely seen. The novel viral strain, SARS-CoV-2, is highly contagious and hence spreads easily when humans come in close contact. This motivates a closer *ad rem* look at the inherent dynamics of the spread at a micro-scale which can help assess its multi-fold ramifications on the cultural, economic and health infrastructures. Using COVID-19 individual-level data and a contact network framework in the initial stages of the pandemic in India, we attempted to answer some critical questions to inform a phased approach to dealing with the pandemic by appropriate resource prioritization and allocation.

At a fundamental level, epidemics spread over a human to human network — where carriers spread the contagion by coming in contact with other people. A simple illustration is available in Figure 1, where the population-level network consists of 100 individuals (nodes), and they are potentially connected among themselves via edges that are meant to simulate human to human interactions. The blue, red and green nodes indicate individuals in the susceptible (“at risk”), infected and removed (recovered or deceased) states, respectively. A transition from the susceptible to the infected state may happen if a susceptible node is connected to an infected node, akin to an uninfected individual coming in contact with an already infected individual. The changing composition of the nodes over time exemplify the typical spreading mechanism of an infectious disease like COVID-19. Such networks are established through various channels such as social and professional interactions, transport and travel or simply any form of shared physical proximity. As shown in Figure 1, where nodes represent hypothetical individuals, the dynamics of spread of the disease is exhibited across time intervals. Established models for epidemic spread, which are further discussed in Section 2, assume random and homogeneous mixing between individuals, which might not be realistic since the general population has defined organizational units (geographical, social etc.) and individuals tend to interact within their sphere of influence. These “links” between individuals form a network and contagions spread along the infection paths wired along these networks. Containment procedures such as lockdowns, social distancing, smart quarantines effectively break this human to human “link” to disconnect the infected network, and hence confine and reduce the spread. Subsequently, using contact networks developed with high geo-specificity and resolution, one can learn specific spread mechanisms and inform precise action planning for mitigation and building health care capacities.

**Figure 1:**
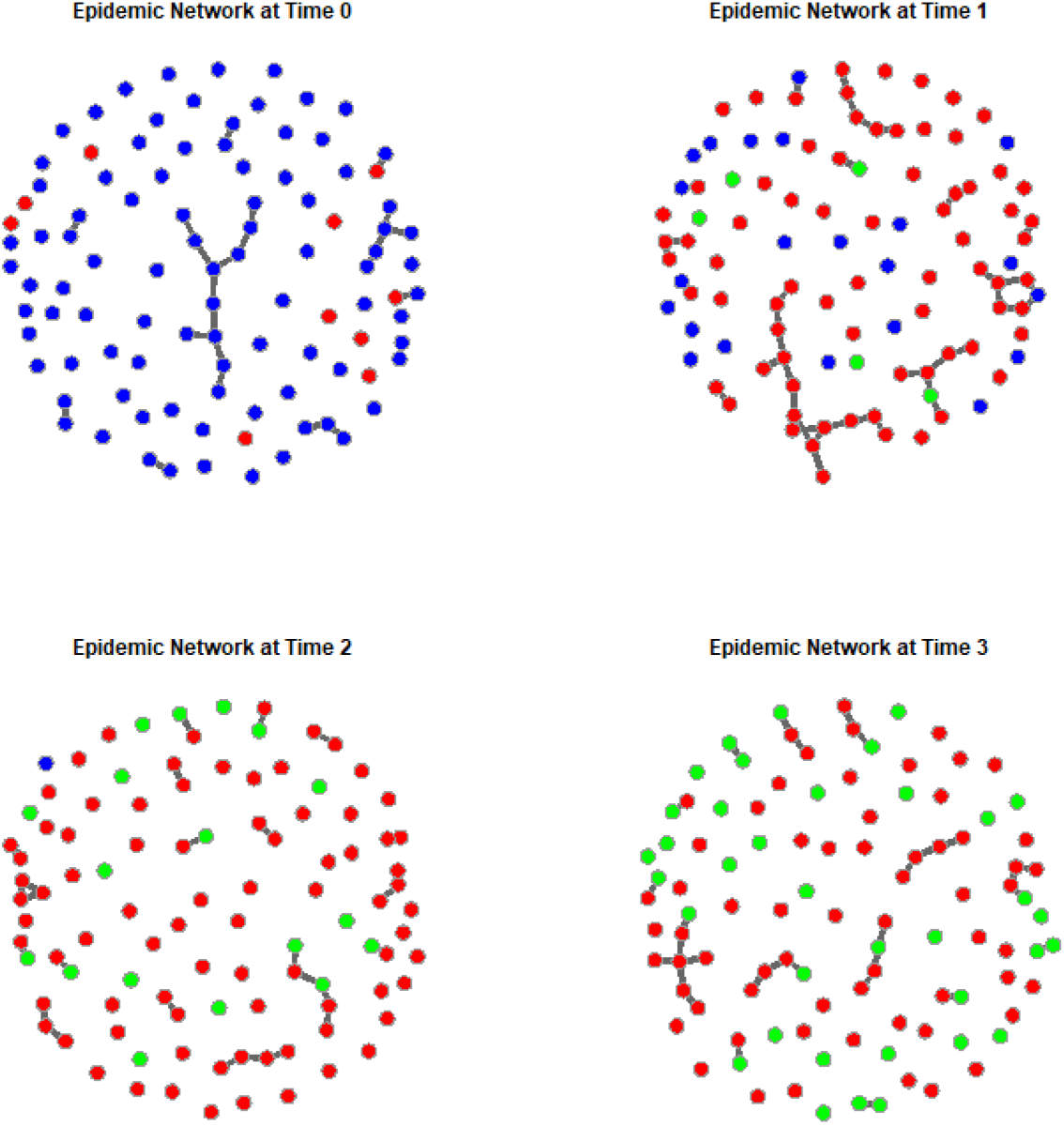
A simple illustration of an epidemic network evolution over time, simulated from a network Susceptible-Infected-Recovered (SIR) model, where the nodes are individual patients: blue are susceptible, red are infected, and green are recovered individuals. One time-step in the figure represents 25 units of time in the original scale of transmissions.

### The India context

India is a large democracy with a population of >1.3 billion^1^. The federal union of India consists of 28 states and 8 union territories, majority of which are contiguous^2^. The first confirmed COVID-19 case in India was reported on January 30, 2020^3^. As per the data released by the Ministry of Health and Family Welfare, Government of India, till the end of March 7, 2021, the cumulative number of confirmed cases in India was more than 11.2 millions, with a little more than 180,000 out of those cases still being active and more than 157,000 of the confirmed cases having been deceased^4^. The large population size and high population density of India rendered it to be on a potentially vulnerable ground from day one. While the eventual fatality rates are relatively low (1.45%, compared to a global fatality rate of 2.2%)^5^, a large population consisting of several significantly sized sub-populations with known co-morbidities had spelled potential troubles for India from the very beginning (Gupta et al., 2011; Arumugam et al., 2020). At the time of the genesis of this project, in early March 2020, the first set of hot-spot Indian states like Kerala, Maharashtra, and Delhi were experiencing rising counts of cases while a number of other states were only beginning to have a surge in their relatively lower case numbers till date.

To address some of these challenges, we formed a study group consisting of researchers from India and United States with the goals to use regional-level data to forecast the number of infections, identify potential hot-spots and recognize how this delineation could potentially inform a phased approach for deploying healthcare capacity across the different states, in the early stages of the pandemic. We utilized individual-level (location-informed) crowd-sourced data from publicly available databases for a total of 34 states and union territories of India across the dates of January 30 to April 15, 2020, covering a total of around 12,000 confirmed cases. Due to governmental restrictions, the individual-level data with location information were not updated after April 27, 2020, and therefore our network-based analyses could not be performed for the later stages of the pandemic in India. A sample location-based network covering the data used is presented in Figure 2. This figure presents all the individual COVID-19 infected patients who are included in the data used for our analyses, with their location information being used to plot them on the map. More details about the data are discussed in Section 4.

**Figure 2:**
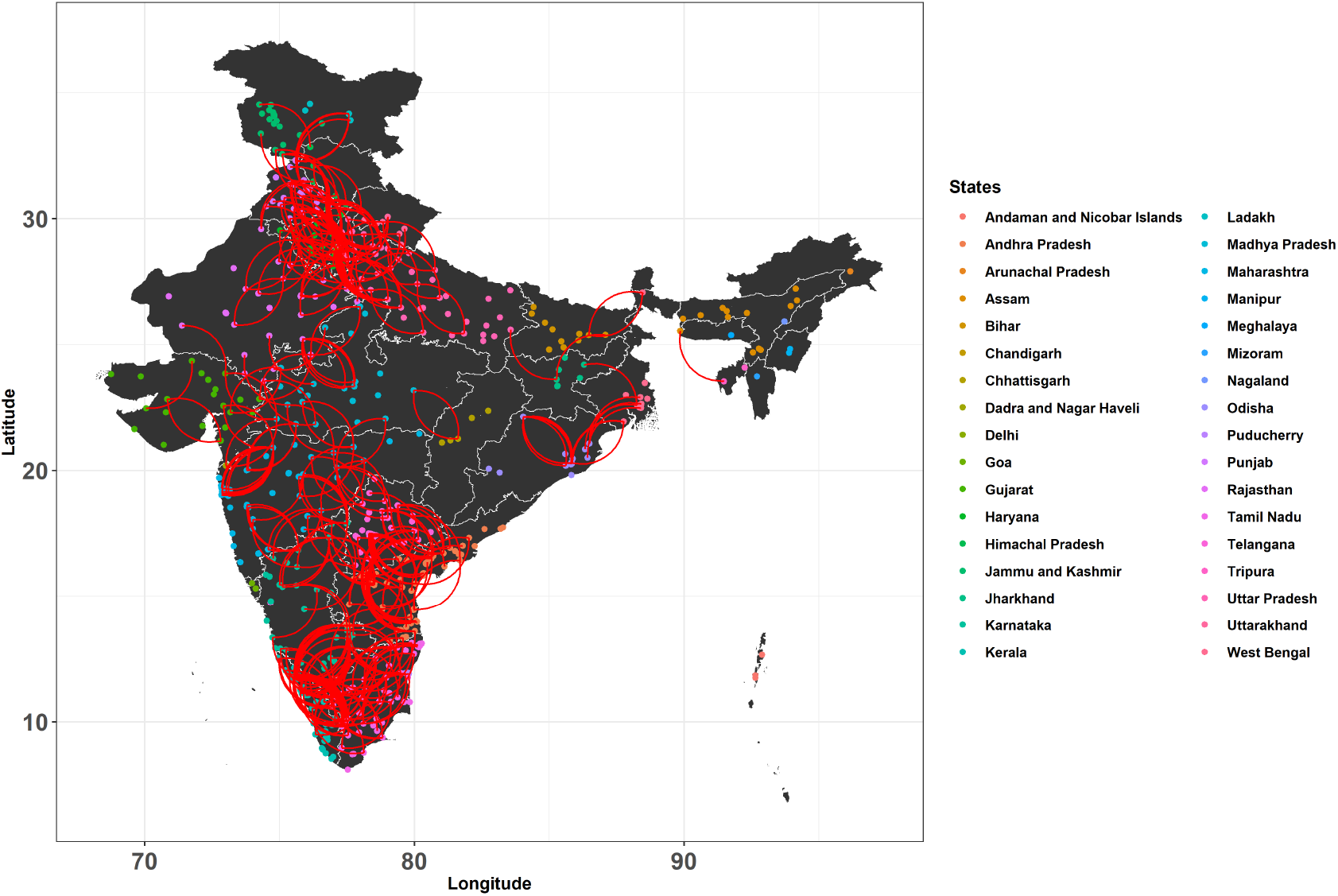
Network of patients infected with COVID-19 across Indian states and union territories, with each dot representing one patient and the color of the dot representing the state in which the patient was first detected and confirmed as a case. Edges are defined by whether two patients are located within a band of two degrees of latitude and longitude of one another. We have removed the edges between nodes that are too close for visual clarity.

In this work, we have gone beyond standard compartmental models for epidemiological modeling via incorporating an underlying network structure among the individuals, respecting their spatial distribution, along with taking into account the population density of respective regions. Besides giving predictions on the number of cases, the network-based analyses for different states and geographical regions provided vital clues for isolation and mitigation strategies for containment of the disease. Specifically, the harsh but necessary strategies like a complete lockdown in the earlier stages of the pandemic, and continued advice on social distancing practices were scientifically validated via our study. We also had several local and national media cover our work. The interactive Shiny application also helped immensely in making demonstrations to the concerned policy makers (e.g. Government officials) and researchers (e.g. public health foundations and institutes) in various Indian states.

The structure of the paper is as follows. In Section 2, we present a brief review of compartmental models in epidemiology, followed by extension of these models in case of network-based inference in Section 3. In Section 4, we present our case study for India, where we describe the available data sets, followed by network-based forecasts for different states. We also present healthcare forecasting in India to assess the healthcare system in place and provide necessary projections for beefing up the infrastructure. We also describe the Shiny application that we have developed in this context. Finally, in Section 5, we provide some discussions on our work including limitations of the study and some potential future directions.

## 2 Compartmental models in epidemiology

One of the widely studied procedures for modeling epidemiological data is using compartmental models (Kermack and McKendrick, 1927). In these class of models, the population is classified into several classes, or compartments, based on the stage of the infection that is affecting them. These models are primarily governed by a system of ordinary differential equations that take into account the time-discoursed infection status of the population compartments. The simplest of such classifications has two population groups, namely, *susceptible*, and *infected*. The corresponding epidemiological models, like the Susceptible-Infected (SI) model or the Susceptible-Infected-Susceptible (SIS) model, take only the aforementioned compartments into account to model the infection dynamics. For a particular disease or infection, the susceptible compartment comprises of those people in the population who are non-infected, but have a potential to catch the disease. On the other hand, the infected compartment comprises of those who have the infection at that given point of time and with a potential to infect others. We now briefly discuss the basic compartmental models alongside explaining the various other compartments and the system of ordinary differential equations (ODEs) governing them.

### 2.1 SI and SIS models

As mentioned earlier, the SI model in the simplest of the compartmental models in epidemiological modeling. This model assumes that all the members of a given population are susceptible to an infection. In the event that a susceptible individual gets infected, the status of that individual changes to being infectious, and that status is retained throughout their life. One supreme example of such disease is the Cytomegalovirus (CMV), a genus that includes the species *Human betaherpesvirus 5* or *HHV-5* that affects humans. This infection has the ability to remain latent within the human host (Landolfo et al., 2003).

We denote the total population size by *N*, and the number of susceptible and infected individuals at time *t* by *S*(*t*) and *I*(*t*) respectively, so that *N* = *S*(*t*) + *I*(*t*). The SI model dynamics is given as

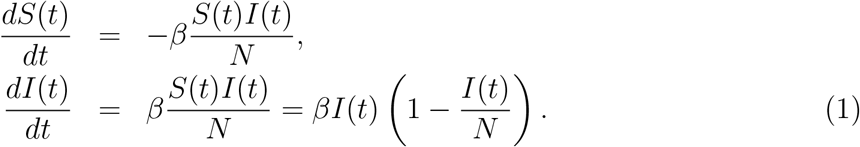

The parameter *β* denotes the infectious rate, defined as the probability of an infectious individual transmitting the infection to a susceptible individual. Denoting the fraction of the population that are susceptible or infected as *s*(*t*) = *S*(*t*)/*N, i*(*t*) = *I*(*t*)/*N* respectively, the SI model has an explicit solution given by

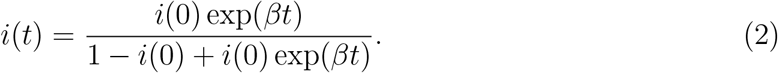

If we monitor the growth in the number of infections with time, the above solution gives that for *t* → ∞, *i*(*t*) → 1, so that eventually the entire population becomes infected.

For diseases where an infected individual becomes susceptible to be re-infected, the SIS model is used. Such diseases include infections caused by rhinoviruses (common cold) (Jacobs et al., 2013), and sexually transmitted bacterial diseases like Chlamydia and Gonorrhea (Hosenfeld et al., 2009). The disease dynamics are given as

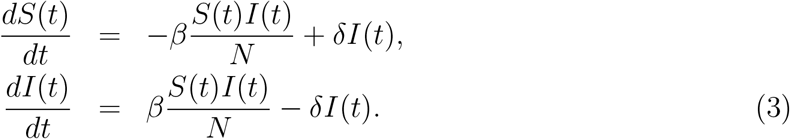

Observe that apart from the previously defined infectious rate *β*, there is another parameter, *δ*, that defines the rate at which infectious individuals return to the state of being susceptible. Explicit solution to the above model is available as well, which is given by

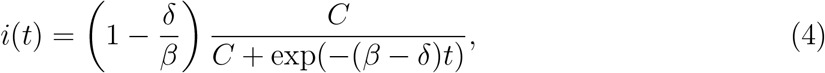

where *C* = *βi*(0)/(*β* — *δ* − *βi*(0)). For *β > δ*, we get a logistic growth curve similar to the SI model. However, unlike the basic SI model, in this case the entire population is not infected, and the infectious fraction approaches (1 — *β*/*δ*) as *t* → ∞. This is referred to as the *endemic state* of the disease, where a certain fraction of the population always stay infected. If *β < δ*, the infections decay to zero exponentially. Hence the threshold value of *β*/*δ* acts as the endemic threshold which governs whether the disease will become an endemic or will eventually die out.

### 2.2 SIR and SIRS models

When an infected individual gains total immunity against the disease, they enter the compartment of being *recovered*, without any chance of getting re-infected, possibly via vaccination. In such cases, the Susceptible-Infected-Recovered (SIR) model is used. Examples include airborne diseases like mumps, measles, rubella, and whooping cough (pertussis). In cases of infections like seasonal flu, where the immunity diminishes with time, a recovered individual becomes prone to re-infection, in which case the SIR model can be extended to the SIRS model. We discuss the dynamics below. Note that with the introduction of a new compartment of recovered individuals (denoted by *R*(*t*)), we need to specify the *recovery rate* (*γ*). The SIR model is given by

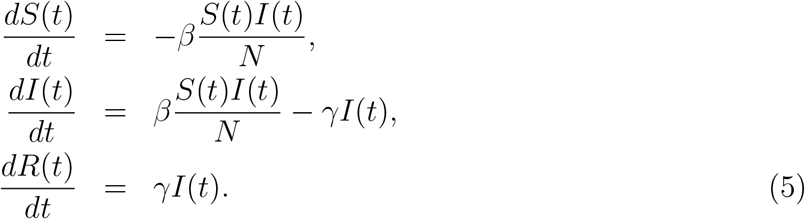

The probability distribution of the duration of infection *τ*, which is defined as the length of time an infected individual remains infected before entering the recovered compartment, is given by an exponential distribution with mean *γ*. This gives the mean infectious time as 1*/γ*. The solution to the above model does not have an explicit form, and simulation based procedures are used to arrive at the solution. It is interesting to explore the size of the outbreak *r*_*∞*_, given by lim_*t→∞*_ *r*(*t*). It satisfies the expression 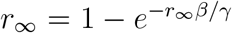. Notice that if *β/γ* ↓ 1, then the size of the epidemic goes to zero, and dies out if *β < γ*. The transition that happens where *β* = *γ* is referred to as the *epidemic transition*. In this regard, we can see the role that is commanded by the ratio *β/γ*. To explore it with more depth, we need to introduce the concept of *basic reproduction number*, denoted by *R*_0_. *R*_0_ is defined as the average number of secondary infections produced by an infected individual during the latter’s infectious period. In case of the SIR model, this can be calculated as *R*_0_ = *β* E(*τ*), which is given by

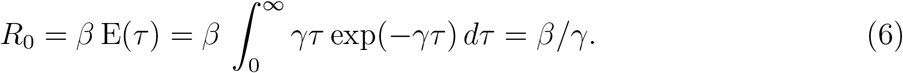

Hence, the disease remains in the epidemic state if *R*_0_ > 1, and ceases to be an epidemic when *R*_0_ drops below unity. For a recovered individual being susceptible again, we need a new parameter, known as the *re-infection rate* (*ξ*), that controls the rate at which re-infections can occur. The resulting SIRS model dynamics is given by

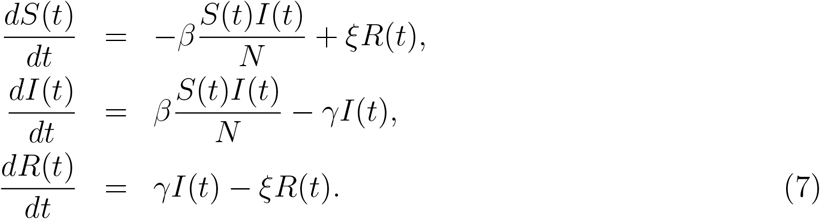

### 2.3 SEIR model

Incubation periods play a vital role in disease dynamics in several infectious diseases. In cases like influenza-type infections, a susceptible individual may get infected, but does not immediately move to the infected compartment owing to latency in the infection and hence is not immediately infectious (Group, 2006). In such cases, a new compartment is introduced, namely *exposed* (E), which acts as a middle step between the susceptible and infected compartments. This leads to an increased complexity of the SIR model, leading to a SEIR model. The dynamics are given as

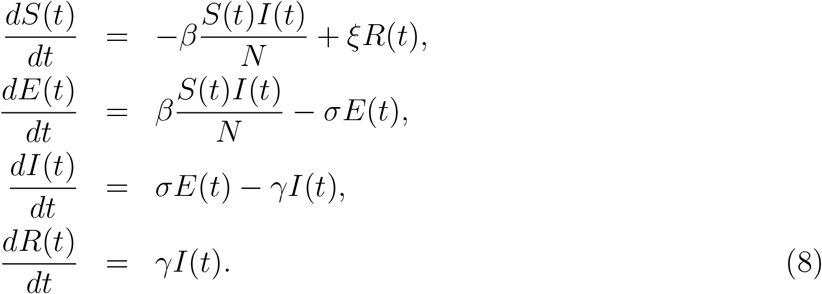

The *incubation rate σ* depends on the average latency period of the infection *L*(= 1*/σ*). This parameter controls the rate at which an infected individual becomes infectious.

Apart from the aspect of compartmentalization, the models discussed above rely heavily upon the assumption of homogeneous mixing, that is, each susceptible individual has equal chance of coming in contact with an infected individual, and hence having the potential to get infected. However, this assumption is very strong, and heavily depends on the nature of infection spread. For example, in case of SARS-CoV-2, the primary mode of spread is through respiratory droplets having a high volume of viral load. Aerosol transmission, though has a low viral load, contributes to infection spread. Other modes include fomites (surface transmission) and airborne transmission. In the latter case, recent research findings show that the virus can stay in the air for as long as 16 hours (Fears et al., 2020). For more details on modes of transmission, we refer the readers to the scientific brief provided by the World Health Organization (WHO) titled ‘Transmission of SARS-CoV-2: implications for infection prevention precautions’^6^. Thus, the infection can only spread via a contact network. Hence it is more appropriate to define the compartmental models over a network of contacts, and explore the network topology to model the spread of the disease. In the next section, we shall discuss epidemiological modeling over a network, that is precisely aimed to capture these phenomena.

## 3 Network-based epidemic models

In mathematical terms, a network is represented by a graph *G* = (*V, E*), where *V* denotes the set of nodes or vertices and *E* denotes the set of edges connecting the nodes. We shall work with undirected edges, where there is no particular sense of directionality in the edges between any pair of nodes in *V*. The adjacency matrix corresponding to *G* is given by a binary matrix *A*, where the (*i, j*)-th element of *A* is given by *A*_*ij*_ = 1 if (*i, j*) ∈ *E*, and 0 otherwise. The degree *d*_*i*_ of the node *i* is defined as the number of edges having node *i* as one of the end-points. Mathematically, we have, *d*_*i*_ = Σ_*j*_ *A*_*ij*_. The starting point of modeling a disease over a network is to first construct the adjacency matrix corresponding to the network. We briefly present the modeling fundamentals over a network structure; for further details we refer the readers to (Newman, 2002, 2018) and references therein.

To model disease dynamics over a network, we need to model the probabilities of the nodes in the network to be in different disease compartments over time. We shall discuss the dynamics of the network-based SIR model in this context. Other related models (like the SI, SIS, SEIR models) over a network are defined similarly. Let us define:

1. *s*_*i*_(*t*): probability that node *i* is in the susceptible state at time *t*,
2. *x*_*i*_(*t*): probability that node *i* is in the infected state at time *t*, and,
3. *r*_*i*_(*t*): probability that node *i* is in the recovered/deceased state at time *t*.

The transmission of infection from an infectious node to a neighboring susceptible node is modeled as a Poisson process with mean *β*, whereas an infected node recovers as a Poisson process with mean *γ*. The disease dynamics of the network-SIR model is then given by

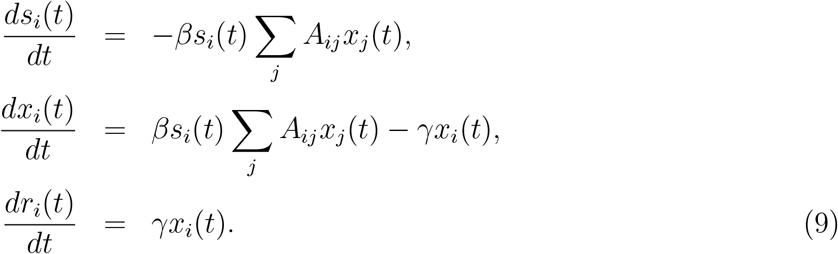

The solution to the above model is approximately given by

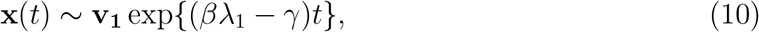

where *λ*_1_ denotes the maximum eigenvalue of *A* and **v**_**1**_ is the corresponding eigenvector. Notice that the underlying network plays an important role in driving the dynamics of the spread of the infection. For a very dense network, with each node having a high degree, the value of *λ*_1_ will be large, so that even for a small value of *β*, the disease can spread exponentially fast. On the other hand, for a sparse network, the maximum eigenvalue *λ*_1_ would be small, so that, for the disease to spread, the infection rate *β* must be very large and the recovery rate *γ* must be very small. Hence it is difficult for the infection to spread in sparse networks. This phenomenon illustrates the scientific principle behind isolation strategies and lockdowns as mitigation procedures in the early stages of a pandemic to curtail the spread of the disease in a given population.

For an arbitrary network with a given degree distribution over the vertex degree *d*, the basic reproduction number *R*_0_ in case of the network SIR model can also be explicitly calculated as,

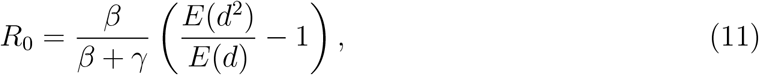

where the expectation is taken with respect to the degree distribution of the network (Andersson, 1998). For a homogeneous network where variability in the degree distribution is low, the reproduction number is low as well. In contrast, for high levels of degree heterogeneity, there are increased chances of ‘super-spreaders’ in the network that can infect a large number of nodes quickly, so that the disease spread becomes more pronounced. This makes the usage of network models more effective, in the sense that if we can trace and isolate these ‘hub’ nodes, then the degree heterogeneity is reduced via dropping of related edges in the network, thus leading to a drop in *R*_0_. This makes network-based modeling especially appealing to model large-scale pandemics such as COVID-19. We now discuss the exact network construction, estimation and parameter choices we used in our setting.

### 3.1 Network construction

Constructing a network of individuals is a big challenge in the sense that micro-level data has to be accessed in order to identify possible contacts and interactions. These include commutation networks, interactions in schools, workplace, social gatherings, to name a few. While the behavior of the disease spread in case of well-defined or simulated networks (for example, Erdos-Renyi graphs or scale-free networks) is mostly known, real-world networks may not exhibit such known structures and would vastly vary on a spatio-temporal basis; see, for example, Masuda and Holme (2013); Karsai et al. (2011); Barabási (2016). Hence we propose to use a spatial network incorporating the geographical location of known infections in the early stages of the pandemic, as we had access to this in our datasets. Formally, the initial network *G* = (*V, E*) is constructed as *V* = *V*_*S*_ ∪ *V*_*I*_, where *V*_*I*_ = set of all infected individuals available in the data, and *V*_*S*_ is the set of susceptible individuals such that |*V*_*S*_| = −*N* |*V*_*I*_| − *N*_*R*_, where *N* is the population size and *N*_*R*_ is the total number of recoveries and deaths (removals) included in the data. | · | Here denotes the cardinality of a finite set. The locations of the added susceptible nodes are initialized by assuming that the expected total number of nodes in any sub-region is proportional to the population density in that sub-region. The nodes *i, j* ∈ *V* are then connected by an edge if *d*(*i, j*) > *δ*, where *d*(*i, j*) indicates the geographical distance between the nodes and *δ* is some user-specified threshold. We present the details about the India-specific choice of this threshold in the next section.

### 3.2 Parameter choices

With the initial network as constructed before, the next step demands selecting the model parameters that would govern the laws of the network evolution and the growth of the pandemic. We note that having fixed the degree distribution, there are two free parameters for a network SIR model – the transmission rate from S to I (*β*), and that from I to R (*γ*). However, more interpretable one-to-one transformations of these parameters are available, namely, the basic reproduction number (average number of susceptible people infected by one infected person - *R*_0_, as expressed in Equation 11), and the average time spent in state I (*T*_*γ*_ = 1*/γ*). For our analyses, we used relevant literature on India to choose ideal values for these parameter that would represent different levels of intensities for the spread of the pandemic. A detailed discussion on these choices are available in Section 4.

### 3.3 Simulation procedure

With the constructed network and the specified values for the two free parameters, we use existing R packages to simulate the growth of this network for a duration of choice. The illustrative simulation in Figure 1 and our data-based simulations described in the next section are performed using the R packages EpiModel (Jenness et al., 2018) and EpiDynamics (Santos Baquero and Silveira Marques, 2020). For the real data-based simulations, a single simulation cycle would yield a time series of compartment memberships as (*S*(*t*), *I*(*t*), *R*(*t*)), *t* ∈ 1,…, *T*_0_, where the initial time point is understood to be time 0, the final time point till which the simulation is performed is denoted by *T*_0_, and *X*(*t*) indicates the number of nodes at state *X* at the given time point. This procedure is then performed M times, denoting the predicted membership for the X compartment at time t from simulation j as *X*^*j*^(*t*), *j* ∈ 1,…, *M, t* ∈ 1,…, *T*_0_. The final predicted count for compartment X at time t is then computed as 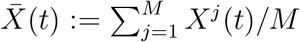. Information on choices of *T*_0_ and *M* for the case study on India are available in the next section. The entire procedure is outlined in Algorithm 1.

#### Algorithm 1: Simulation procedure for network-SIR models

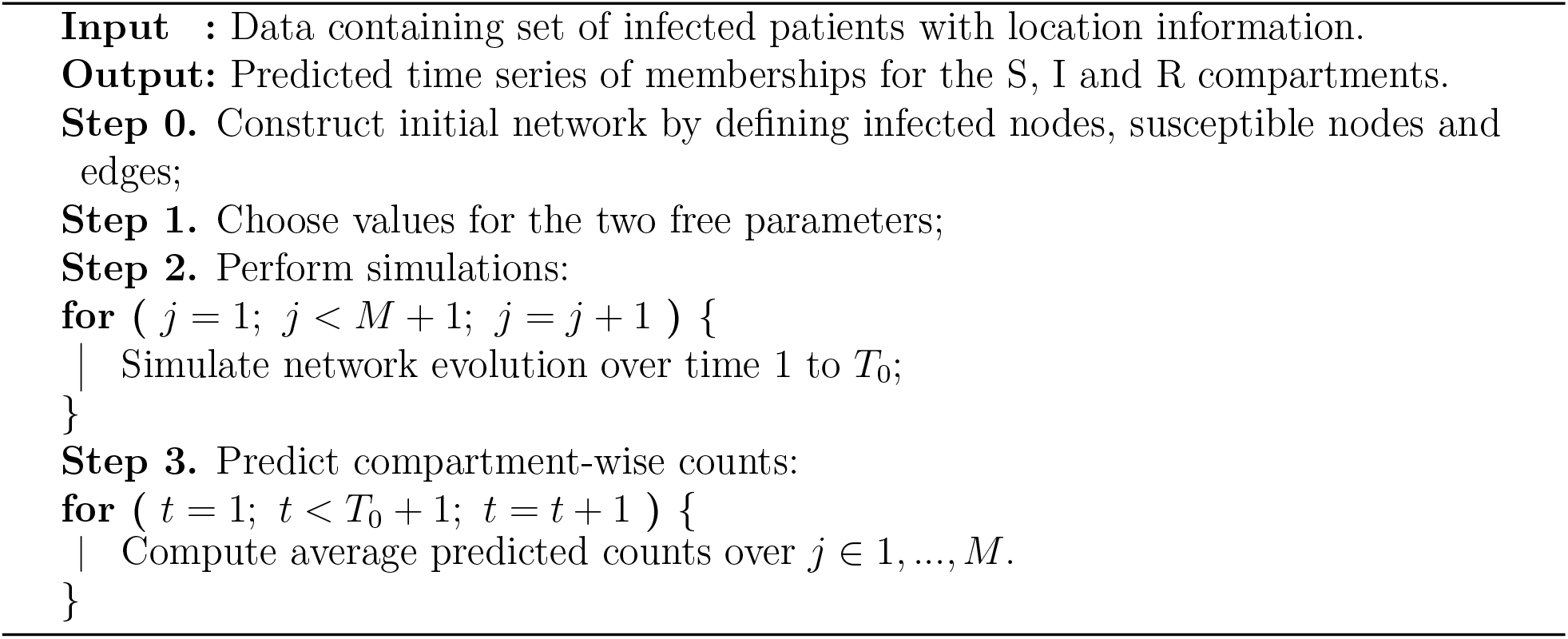

## 4 Case study for India

### 4.1 Data description

Individual-level, location-informed COVID-19 data for Indian states and union territories were obtained from the Kaggle database^7^. The data freeze used for our analyses covered 34 Indian states and union territories having at least one confirmed COVID-19 case within January 30 to April 15, 2020, resulting in a total of approximately 12,000 confirmed cases. State-wise distribution of these cases is available in Table 1, and a location-based visualization across India is available in Figure 2. At the time of this data being obtained, India had just finished the first stage of nation-wide lockdown (25 March to 14 April) and was at the beginning of the second phase of it. Data on state-level availability of hospital beds were obtained from the Ministry of Health and Family Welfare, Government of India press release^8^. The primary analytical tasks were geared towards answering two major questions: forecasting the number of infected cases on a monthly basis for each state in India, identifying potential hot-spots and recognizing how this delineation could inform a phased approach for deploying healthcare capacity across the different states.

**Table 1:** Number of confirmed COVID-19 cases by states and union territories of India as of April 15, 2020 as used for the network SIR analyses.

| State | Confirmed Cases |
| --- | --- |
| Andaman and Nicobar Islands | 11 |
| Andhra Pradesh | 473 |
| Arunachal Pradesh | 1 |
| Assam | 31 |
| Bihar | 66 |
| Chandigarh | 21 |
| Chhattisgarh | 33 |
| Dadra and Nagar Haveli | 1 |
| Delhi | 1510 |
| Goa | 7 |
| Gujarat | 650 |
| Haryana | 198 |
| Himachal Pradesh | 32 |
| Jammu and Kashmir | 278 |
| Jharkhand | 27 |
| Karnataka | 260 |
| Kerala | 386 |
| Ladakh | 17 |
| Madhya Pradesh | 614 |
| Maharashtra | 2455 |
| Manipur | 2 |
| Meghalaya | 1 |
| Mizoram | 1 |
| Nagaland | 1 |
| Odisha | 60 |
| Puducherry | 7 |
| Punjab | 184 |
| Rajasthan | 969 |
| Tamil Nadu | 1204 |
| Telangana | 592 |
| Tripura | 2 |
| Uttar Pradesh | 660 |
| Uttarakhand | 37 |
| West Bengal | 190 |

### 4.2 Network-based forecasts

As outlined in the previous section, for the state-specific forecasts, we used a network-based SIR model. First, we created a national-level network of the infected patients, with the patients as nodes, and two nodes being connected if their location of detection are within both 2 degrees of latitude and longitude of one another. Note that this is equivalent to choosing a value of *δ* as introduced in the previous section. Next, for each state, we extracted the subset of this network falling in that state (determined by the location of detection of the nodes), and that became the initial infected network of the state. The complete initial network was constructed by adding additional susceptible nodes so that the number of nodes matched the population of the state and by assuming zero removed nodes at the beginning.

Earlier literature on the Indian COVID-19 data based on count-based SIR models and their modifications indicated that *R*_0_ = 2 was a good India-specific estimate, and given the healthcare resources and infrastructure of India, *γ* = 0.1 i.e. *T*_*γ*_ = 10 days is a sensible number (Pandey et al., 2020; Singh and Adhikari, 2020). Therefore, we used this value of *γ* across all states, and focused on three potential scenarios of particular interest in erms of the *R*_0_, as outlined below.

- **Scenario 1: Low intensity spread:** Assuming strict adherence to containment and mitigation protocols (*R*_0_ = 1.5).
- **Scenario 2: Medium intensity spread:** Assuming sporadic adherence to containment and mitigation protocols (*R*_0_ = 2).
- **Scenario 3: High intensity spread:** Assuming minimal adherence to containment and mitigation protocols (*R*_0_ = 2.5)

More scenarios are available in our shiny app described in the next subsection. Our network-based forecaster predicted the number of infected cases over short-term future (three months from the last date when data are available, that is, *T*_0_ = 90) and the spread over specific geographical regions over potential contact networks for every state in India. Based on these, we tried to answer which states would potentially emerge as hot-spots.

Figures 3-5 show the predicted number of active COVID-19 cases across Indian states and union territories between April 15 to July 1, 2020. The three figures respectively correspond to the three (low, medium, high) intensity scenarios as described before. In each figure, we present the predicted time series of infected cases per million people for each state during April 16, 2020 to July 1, 2020. The reason behind plotting the prevalence per million population instead of the raw predicted case counts was that the states and union territories of India have extremely varied population sizes, and without adjusting for that, the raw predicted case counts would tend to emphasize upon the larger states, potentially masking some relatively smaller regions demanding more attention for better containment of the pandemic. We also report the short-term prediction performances under the three scenarios as of May 31, 2020, in Table 2.

**Table 2:** Comparison of observed and predicted cumulative COVID-19 case counts as of May 31, 2020. The second column provides the observed number of cases as of that date. The next three columns provide the network-based predictions under the low (*R*_0_ = 1.5), medium (*R*_0_ = 2.0) and high (*R*_0_ = 2.5) intensity scenarios.

| State | Observed | Low | Medium | High |
| --- | --- | --- | --- | --- |
| Andaman and Nicobar Islands | 33 | 114 | 1195 | 11619 |
| Andhra Pradesh | 3571 | 4938 | 51856 | 530809 |
| Arunachal Pradesh | 4 | 11 | 110 | 1159 |
| Assam | 1340 | 312 | 3432 | 35886 |
| Bihar | 3807 | 728 | 7286 | 76513 |
| Chandigarh | 293 | 219 | 2291 | 22794 |
| Chhattisgarh | 498 | 357 | 3627 | 38164 |
| Delhi | 19844 | 15730 | 159905 | 1345083 |
| Goa | 71 | 72 | 770 | 7986 |
| Gujarat | 16794 | 6829 | 71258 | 726726 |
| Haryana | 2091 | 2078 | 21726 | 223599 |
| Himachal Pradesh | 331 | 336 | 3521 | 36526 |
| Jammu and Kashmir | 2446 | 2909 | 30299 | 299686 |
| Jharkhand | 635 | 296 | 2968 | 31305 |
| Karnataka | 3221 | 2749 | 28592 | 297167 |
| Kerala | 1270 | 4042 | 42292 | 430503 |
| Ladakh | 77 | 177 | 1821 | 16319 |
| Madhya Pradesh | 8089 | 6463 | 67397 | 691479 |
| Maharashtra | 67655 | 25733 | 267675 | 2649000 |
| Manipur | 71 | 19 | 219 | 2318 |
| Meghalaya | 27 | 11 | 112 | 1189 |
| Mizoram | 1 | 10 | 110 | 1157 |
| Nagaland | 43 | 9 | 110 | 1161 |
| Odisha | 1948 | 629 | 6631 | 69299 |
| Puducherry | 70 | 73 | 768 | 7960 |
| Punjab | 2263 | 1942 | 20197 | 208574 |
| Rajasthan | 8831 | 10145 | 105975 | 1071343 |
| Tamil Nadu | 22333 | 12625 | 131596 | 1320651 |
| Telangana | 2698 | 6194 | 64686 | 649082 |
| Tripura | 316 | 22 | 224 | 2321 |
| Uttar Pradesh | 8075 | 6993 | 72531 | 756689 |
| Uttarakhand | 907 | 383 | 4074 | 42372 |
| West Bengal | 5501 | 2008 | 20902 | 218880 |

**Figure 3:**
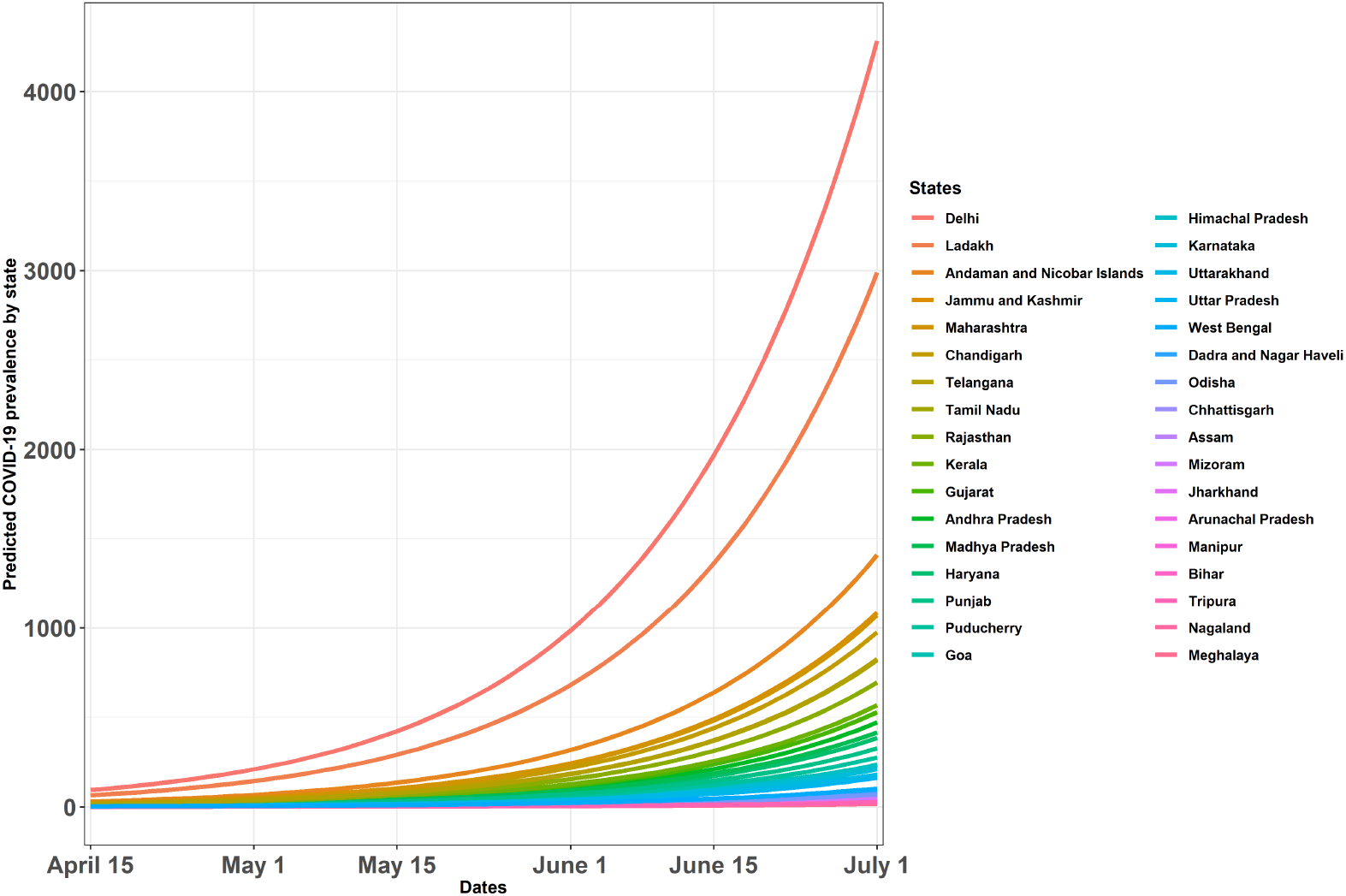
Predicted number of active COVID-19 cases per 1,000,000 people across Indian states and union territories based on network SIR (susceptible-infected-removed) models. The initial network for each state contains the infected individuals, and an additional number of susceptible individuals so that the total size of the network matches the population of each state. Edges are defined by whether two persons are located within a band of two degrees of latitude and longitude of one another. For running the simulations, the reproduction number *R*_0_ was assumed to be 1.5 (low intensity scenario), and the average time at state I was taken to be 10.

**Figure 4:**
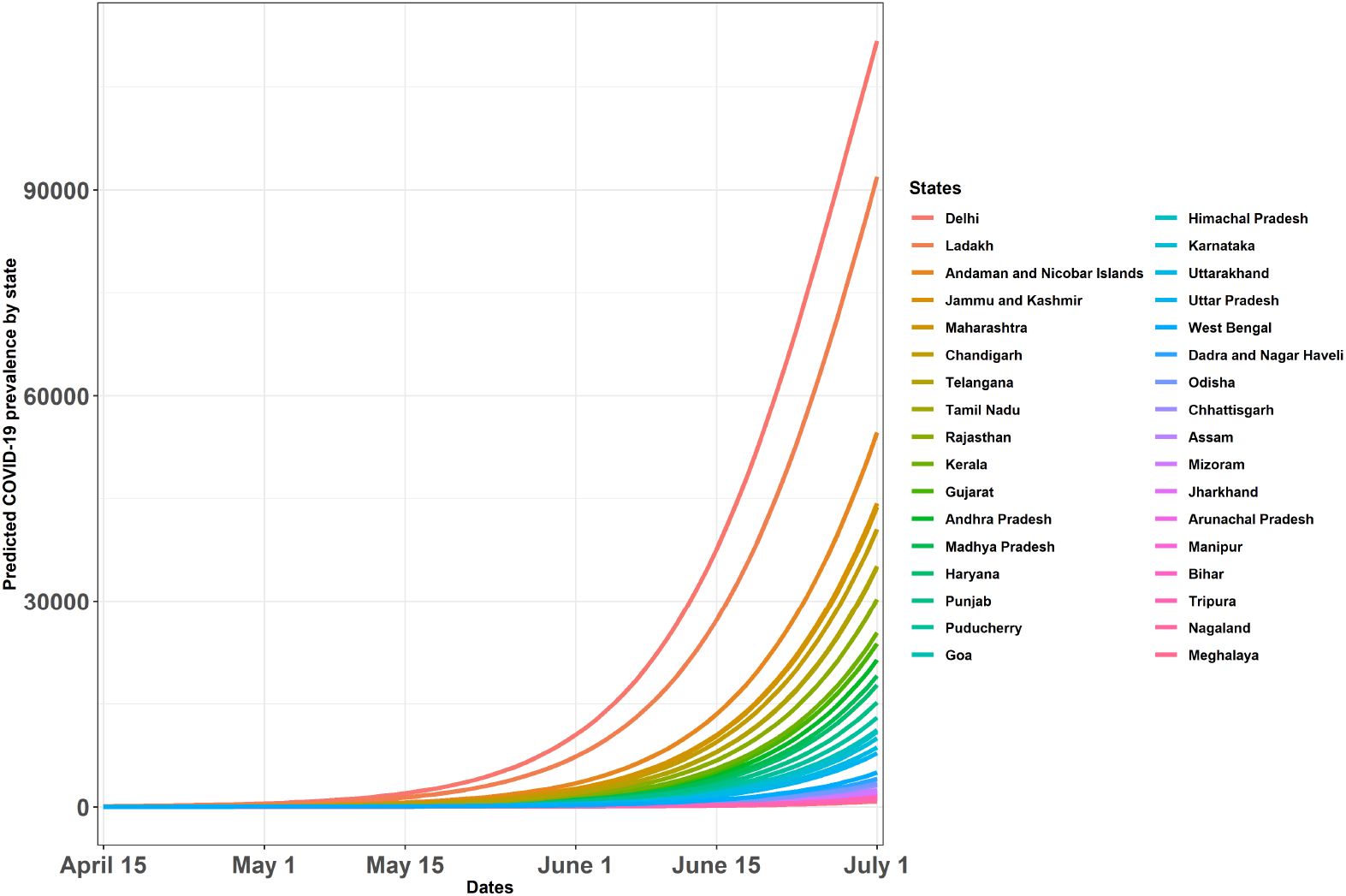
Predicted number of active COVID-19 cases per 1,000,000 people across Indian states and union territories based on network SIR (susceptible-infected-removed) models. The initial network for each state contains the infected individuals, and an additional number of susceptible individuals so that the total size of the network matches the population of each state. Edges are defined by whether two persons are located within a band of two degrees of latitude and longitude of one another. For running the simulations, the reproduction number *R*_0_ was assumed to be 2 (medium intensity scenario), and the average time at state I was taken to be 10.

**Figure 5:**
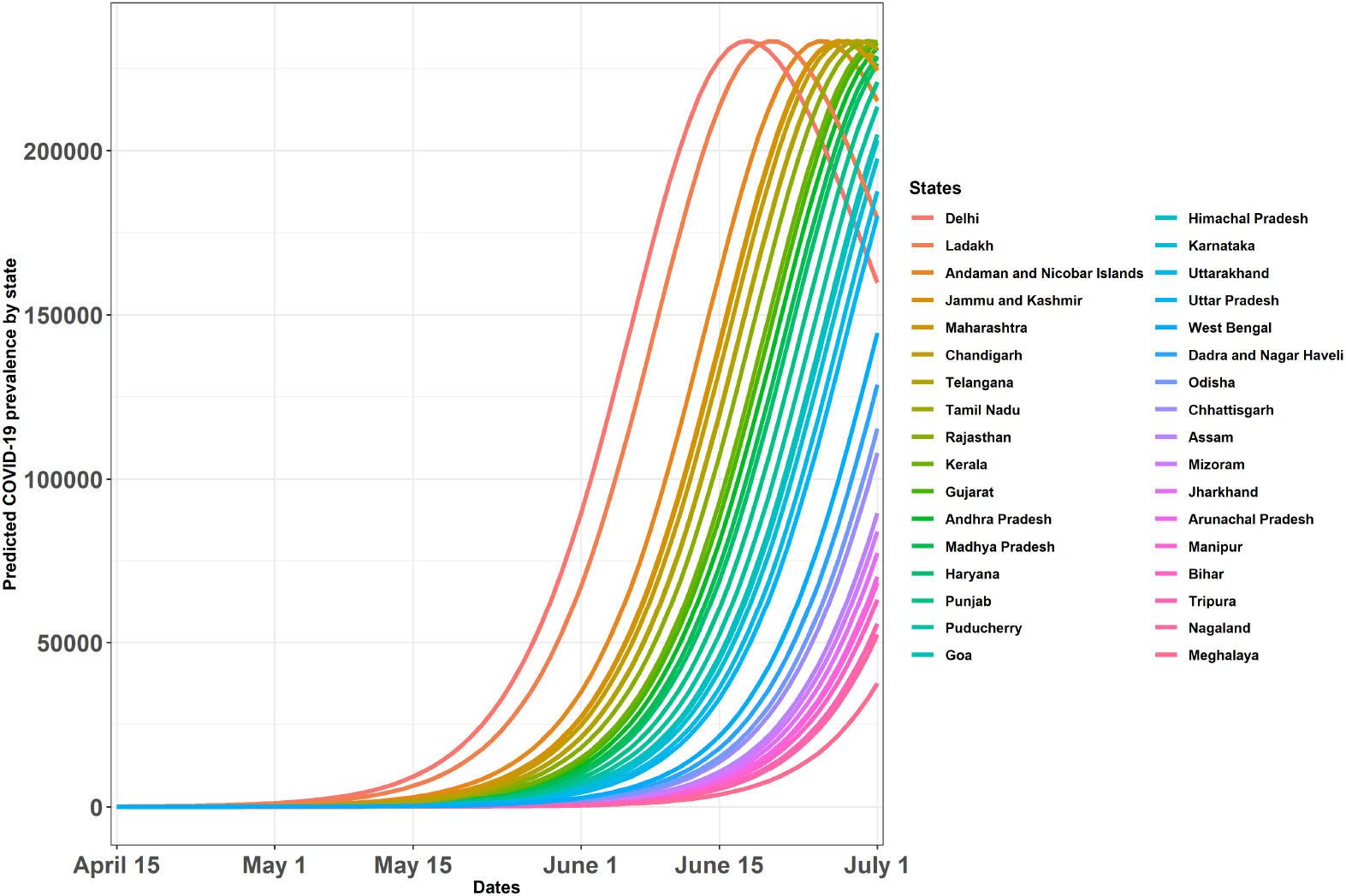
Predicted number of active COVID-19 cases per 1,000,000 people across Indian states and union territories based on network SIR (susceptible-infected-removed) models. The initial network for each state contains the infected individuals, and an additional number of susceptible individuals so that the total size of the network matches the population of each state. Edges are defined by whether two persons are located within a band of two degrees of latitude and longitude of one another. For running the simulations, the reproduction number *R*_0_ was assumed to be 2.5 (high intensity scenario), and the average time at state I was taken to be 10.

Some evident trends emerged. Most states had a significant rise in the number of cases starting around mid-May. There were clear hot-spots emerging in a first cluster of states that included Maharashtra, Delhi, Tamil Nadu and Rajasthan with a second cluster (close behind) emerging in Uttar Pradesh, Gujarat, Madhya Pradesh, Telangana, Andhra Pradesh and Kerala. While a number of these initial hot-spots could be traced to international travel and port of entries, some newly formed hot-spots could potentially be attributed to hub-and-child nodes of contact networks through travels and public gatherings.

While these predicted numbers were in the hundreds over the initial few weeks, they rose multi-fold to thousands to hundred thousands, surging to millions and tens of millions — under the assumed rates, especially under the high intensity scenario. Figures 3-5 show the number of affected cases increasing substantially in late May through June in all three scenarios, where some states were predicted to cross a million of cases by mid-June. Unsurprisingly, states with high population density (e.g. Maharashtra, Rajasthan, Madhya Pradesh and Karnataka) were seen to be extremely susceptible to see a spike in the number of cases. Looking at some of these states case by case, we could interpret how our model was able to capture the ground reality, retrospectively. The next subsection discusses these state-specific patterns in more detail.

### 4.3 State-specific projections

We now present the short-term (six weeks) predictions for May 31, 2020 considering three scenarios - low, medium and high intensity spreads as given by the choice of *R*_0_ = 1.5, 2 and 2.5 respectively. The corresponding predcitions serve as adhoc prediction intervals for the number of infections across different states. The results are presented in Table 2. Almost all the states record infections that are within the range of predicted number of infections. In this regard, we re-iterate that the goal of these predictions were to present vital clues for isolation and mitigation strategies for containment of the disease, as well as increase preparedness to treat patients with infections along with ramping up the overall healthcare system. We now discuss some key policy adoptions taken by different states and regions within the country, where regional network management played a vital role.

Kerala, a southern state in India, had reported the first case in the country on January 30, the patient being a student who had traveled from Wuhan, China. The state, which is a favorite among tourists, witnesses a massive foreign footfall every year. Also, it has one of the highest percentages of expatriates and domestic migrant workers. These served as points of concern, making Kerala one of the most vulnerable states. To tackle this, the state released a public health advisory guideline on January 26, 2020, that directed serving a mandatory isolation period for all people traveling from China. Kerala aggressively put itself on war-footing to combat the upcoming peril by mobilizing its strong healthcare workforce in efforts towards vigorous testing, contact tracing to finer levels, increasing duration of quarantine, and ensuring that the migrant workforce are sheltered and well-fed (Rahim and Chacko, 2020). Kerala initially showed promising results, with the percentage of new cases on the decline, along with a steady increase in recovery rate. However, with the gradual unlockdown procedure and an increased rate of contact, Kerala eventually experienced another spike in terms of daily infections. A hike in daily cases of over 100% was observed in the state after the festival season of *Onam* that was celebrated in the last week of August to the first week of September^9^.

In comparison, Maharashtra, the worst affected state as of the data freeze in mid-April, had registered almost similar number of cases by the end of March, but started performing poorly, primarily owing to insufficient number of testing, tracking and isolation. With its capital Mumbai being the commercial hub of the country, a significant portion of infections in the population were left untraced due to inadequate levels of testing^10^. Another major talking point comes from the Bhilwara district in Rajasthan, once deemed as ‘India’s Italy’ by BBC^11^. Bhilwara had been successful in restricting the infection from spreading in less than 2 weeks from its first reported case, with the last reported case registered on March 31 (during the period of our study). The success story had been attributed to massive testing, extensive tracking and enforcing strict isolation, apart from door-to-door surveys and continued screening of the population in the district (Meghwal et al., 2020). This led to screening a whopping number of 2.2 million people in the district which accounts for 92% of its population. The Bhilwara model serves as a paramount example of how the policy of testing-tracing-isolation-treatment can work wonders in containing the spread of the disease. Odisha had been dynamic as well from the very beginning of the outbreak, starting with locking down its capital Bhubaneshwar from March 12, before the state-wide lockdown from March 24 (Lancet, 2020). The state closely followed the Bhilwara model and had also announced an extension of the lockdown in the state till April 30, even before the announcement of the second phase of country-wide closure. The strong health workforce had synchronized in complete harmony with numerous self-help groups and policy-makers to effectively take benefit from the lockdown.

### 4.4 Comparison with sero-survey findings

Seroprevalence studies are crucial in determining the infection prevalence in a region. These survey based studies check for the presence of antibodies of a disease using blood serum specimens. The Indian Council of Medical Research (ICMR), along with Dept of Health and Family Welfare, Govt. of India and National Center for Disease Control, on May 12, 2020, announced to conduct multiple national level community based sero-surveys across several states in India. The subsequent sero-survey findings are quite interesting. The first serosurvey was conducted in May-June 2020 among adults aged 18 or older across 21 states in the country. Although the seroprevalance was reported to be as low as 0.73% (95% CI: 0.34% - 1.13%) (Murhekar et al., 2020), the infection-to-case ratio was estimated to be between 81.6 and 130.1 infections per reported case of the disease till June 2020. Lockdown relaxations were being adopted across the country in phases after this period, and a second serosurvey exercise was done during August-September 2020. The second serosurvey also included individuals who are 10 years or older. The sero-prevalance was estimated to be 7.1% (95% CI: 6.2% - 8.2%) among adults aged 18 years or older, with estimated 26-32 infections per reported case, thus leading to around 74.3 million infections in the country by mid-August 2020. The detailed findings have been reported in Murhekar et al. (2021), including details on the districts and states included in the survey. Our network-based estimate were 0.13 million, 3.95 million, and 6.32 million on August 12, 2020, the last day of prediction, under the three scenarios, respectively. The observed number of infections falls within this range, but the seroprevalence adjusted estimate is much higher. We note that it is not readily possible to validate the seroprevalance estimates with the network-based estimates owing to the fact that we had constructed the network using the observed cases only. The third round of the ICMR serosurvey conducted during the period of December 2020 - January 2021, reported roughly a three-fold increase in the number of infections since August 2020, with a infection prevalence rate of 21.5% for persons aged 10 years or above. India has a reported number of a little over 10 million infections, thus putting the estimated number of infections to be around 270 million (27 actual cases per reported case). These results have been released by ICMR via a press-conference on February 04, 2021, and are yet to be formally published (as of the time of writing this article). The serosurvey findings thus validate, to an extent, the estimates based on our network-based methods, additionally indicating that managing regional contact networks are crucial along with other best practices such as maintaining basic hygiene practices and wearing of masks.

### 4.5 Healthcare forecasting in India during the COVID-19 pandemic

At a national-level, several steps for containment and mitigation were already put in place (by mid-April 2020) by the government. These included imposition of an extended nationwide lockdown and campaigns to increase awareness about preventive and behavioral measures. Such measures were expected to contribute towards flattening the curve, which was absolutely essential to prevent a situation where the healthcare infrastructure of the country being no longer able to handle the burden. The predictions of our network-based models showed alarming counts for the number of people who could potentially get infected over time. Such a situation could overburden the existing healthcare infrastructure in the country. Specifically, our initial forecasts indicated that the public healthcare facilities might see a huge surge in patients as the majority of the Indian population depends on public healthcare resources. This was the ground situation indeed till the point where the infection growth curve started coming down. Hence, it was important to assess if the infrastructure was ready to handle a huge surge of COVID-19 confirmed cases.

We considered the healthcare capacity in India across different states to analyze the overall availability of hospital beds in India. We obtained data about the hospital beds across the country from a press release^12^ by the Ministry of Health and Family Welfare. Additionally, we obtained the population for each state from the Open Government Data Platform of India^13^. The levels of healthcare resources required will vary based on the different clinical outcomes of the infection as well as the existing availability of healthcare infrastructure. We captured the former aspects using a compartmental epidemiological model based on the classical SEIR model. The infection levels are broadly categorized as mild, severe and critical, and we focus on the medium intensity spread scenario with the reproduction number *R*_0_ = 2 under intervention (for further data and details see our shiny app here). Note that we adapted the SEIR model and the publicly available R shiny app developed by Dr. Alison Hill’s group^14^ to forecast the healthcare capacity. The different parameter values were also chosen based on the guidance provided in this R shiny app (specific parameter values are also provided on our shiny app).

The predictions based on the SEIR model indicated a peak for the number of cases around mid-May. In Figures 6-8 (Panel A), we show the state-wise number of additional hospital beds required (states sorted from the lowest requirement to highest) in a waterfall plot assuming the percentage of prior occupancy at 25%, 50%, and 75%, respectively. Assuming that half of the cases who are hospitalized will need ventilators, we also plotted the requirement of ventilators across these states. These additional hospital bed requirements are also plotted on a map of India for all the states and union territories in Figures 6-8 (Panel B). We saw that many smaller states such as Arunachal Pradesh, Goa, Mizoram and Sikkim, and the union territories had sufficient healthcare infrastructure to deal with a surge under the assumptions considered. Whereas states such as Uttar Pradesh, Bihar, Maharashtra, and West Bengal were forecast to be in desperate need of additional hospital beds and healthcare infrastructure to successfully deal with a pandemic of such scale.

**Figure 6:**
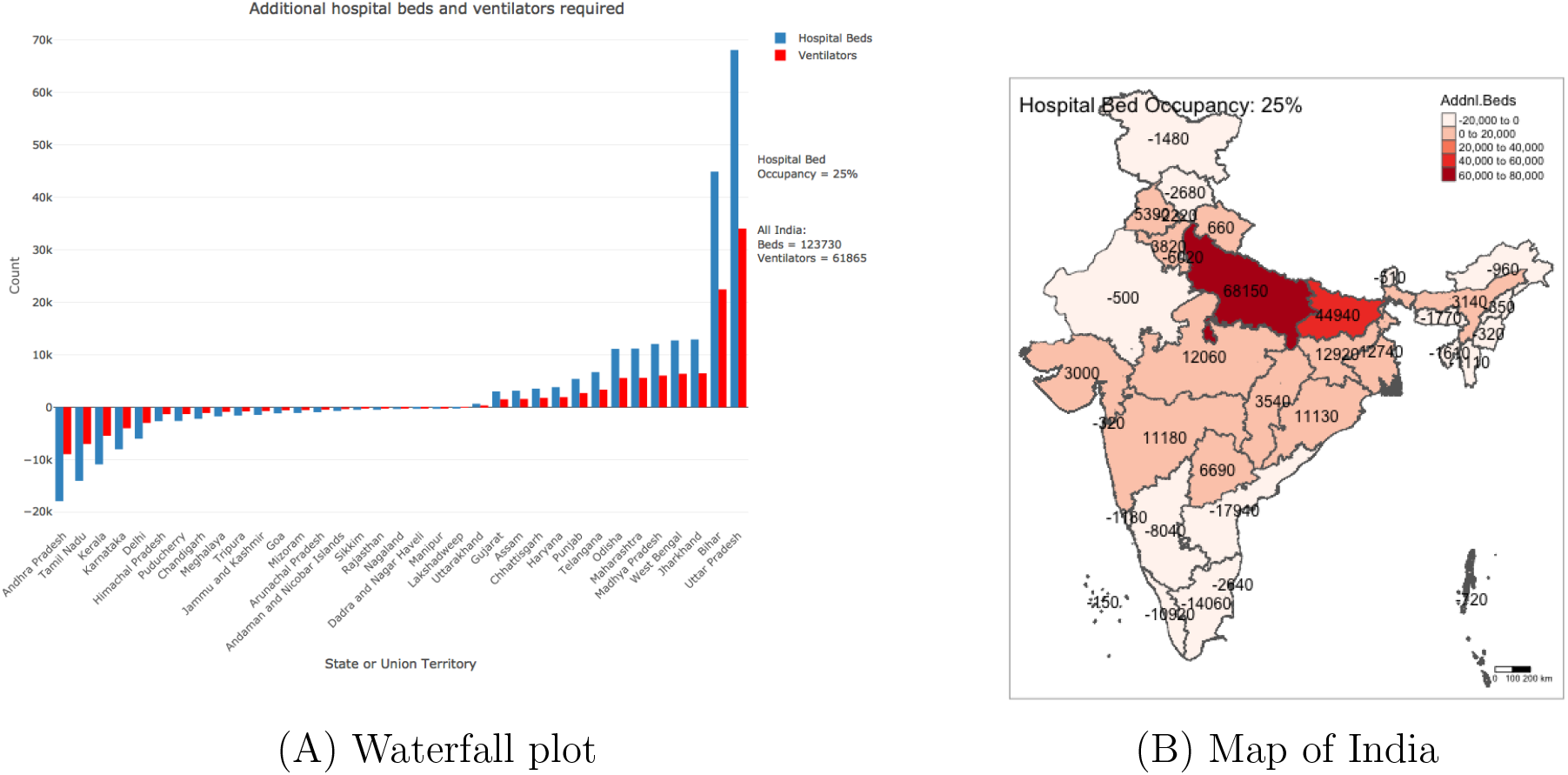
(A) Waterfall plot and (B) map of India showing the shortfall of hospital beds and ventilators across Indian states and union territories on the predicted peak day of incidence of new cases. These estimates are based on a SEIR model with the reproduction number *R*_0_ = 2 under sporadic adherence to containment and mitigation protocols assuming the normal occupancy rate of the hospital beds as 25%.

**Figure 7:**
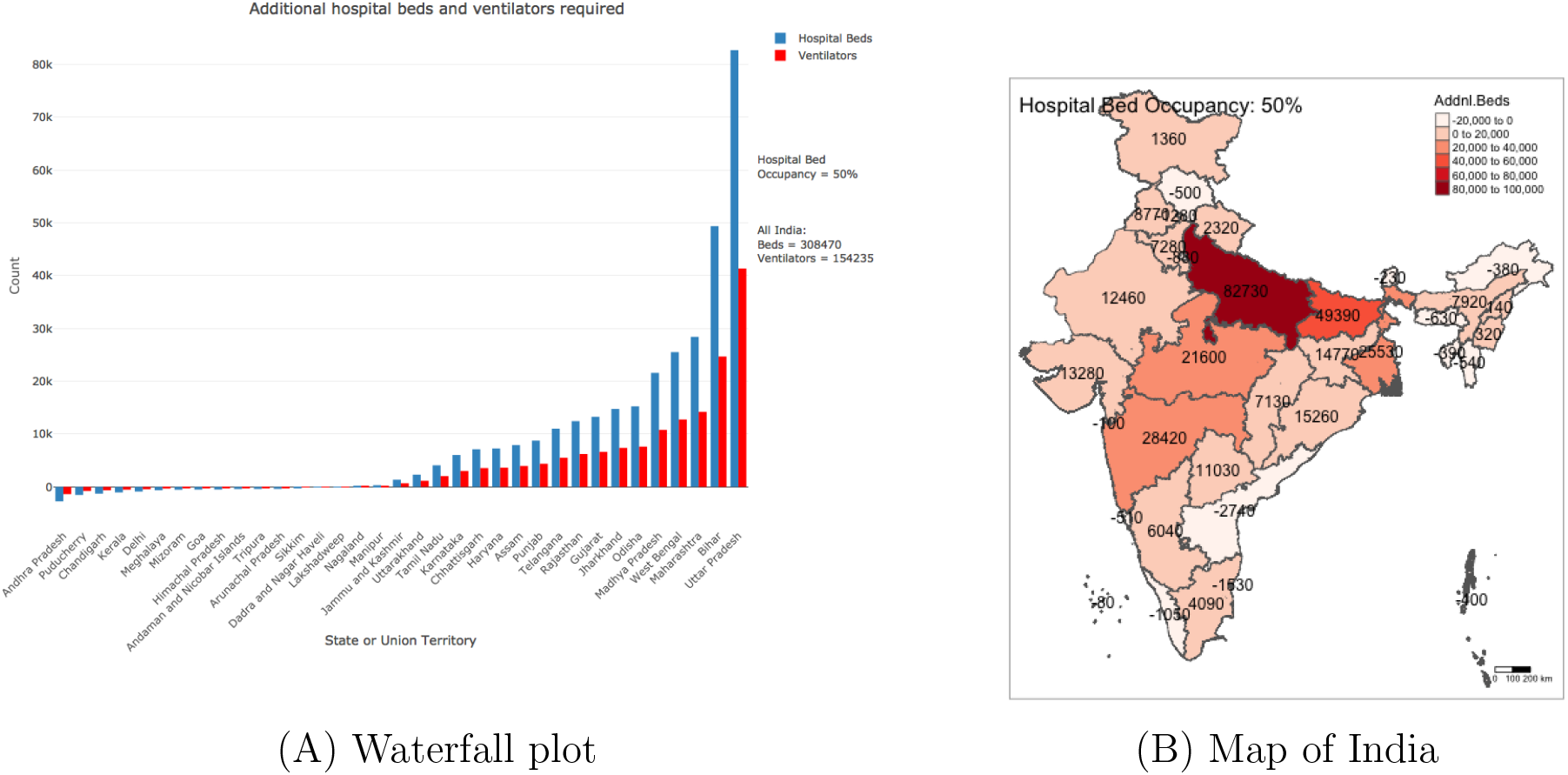
(A) Waterfall plot and (B) map of India showing the shortfall of hospital beds and ventilators across Indian states and union territories on the predicted peak day of incidence of new cases. These estimates are based on a SEIR model with the reproduction number *R*_0_ = 2 under sporadic adherence to containment and mitigation protocols assuming the normal occupancy rate of the hospital beds as 50%.

**Figure 8:**
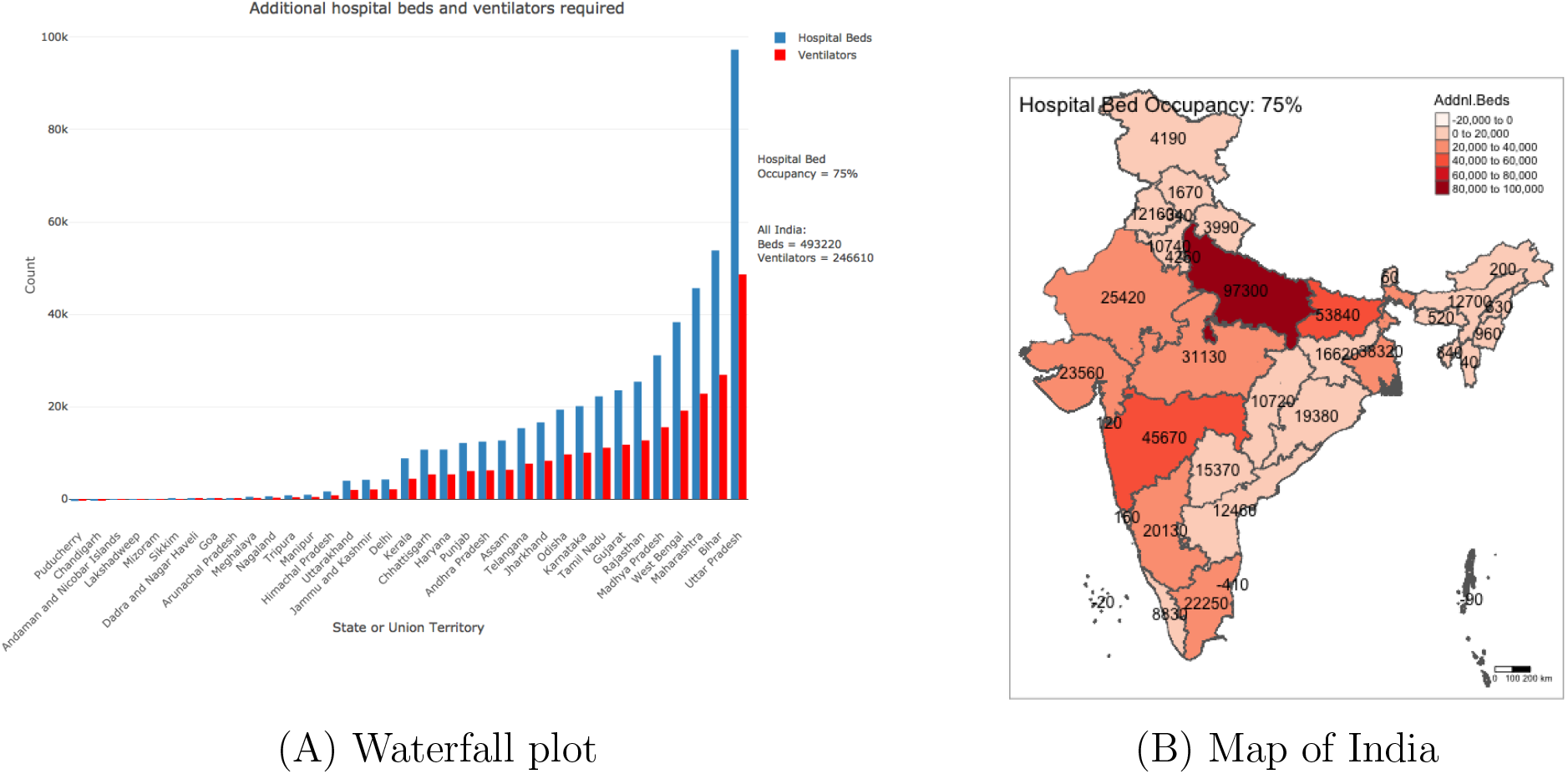
(A) Waterfall plot and (B) map of India showing the shortfall of hospital beds and ventilators across Indian states and union territories on the predicted peak day of incidence of new cases. These estimates are based on a SEIR model with the reproduction number *R*_0_ = 2 under sporadic adherence to containment and mitigation protocols assuming the normal occupancy rate of the hospital beds as 75%.

Our results indicated the immediate need for the administrators to mobilize resources and infrastructure in hotspot areas and acquire the appropriate number of hospital beds (permanent or makeshift), ventilators, personal protective equipment, and the accompanying personnel to support the huge surge that followed ahead.

### 4.6 Shiny dashboard and resource availability

With a goal to making our results more accessible and to allow interested researchers and policymakers alike to visualize the simulated projections under a broader range of scenarios, we had built an R shiny dashboard offering a multitude of functionalities. A representative snapshot of the different functionalities of the dashboard is available at Figure 9. The interactive dashboard attempts to forecast the state-wise number of COVID-19 cases for India over the weeks/months following April 15, 2020 using state-level case data and the network-based SIR compartmental model approach as described before. Further, it offers SEIR model-based simulations to predict the potential shortage in healthcare facilities and infrastructure for specific regions, as outlined in the previous subsection. All the plots are interactive and update themselves based on user inputs for several epidemiological and infrastructural parameters. The user may also hover over the figures for looking at the exact numbers, and click the camera icon on top of each figure to download the plots as png files. The home page of the dashboard sets up the network modeling context with respect to the Indian COVID-19 scenario, using figures akin to Figures 1-2 in this paper. The “Case Predictions across States/UTs: Network SIR” tab outlines the network SIR model briefly, with relevant references for the theoretical backgrounds and some discussions on the parameter choices and simulation procedure. It offers case forecast time series plots similar to those in Figures 3-5 in this paper (for the entire country or for a particular state/union territory) under user-specified values of the basic reproduction number *R*_0_ and the average time spent in infected state *T*_*γ*_. Dynamic bar plots are available for the three intensity scenarios discussed in this paper. The “Healthcare Forecasting: SEIR” tab describes the SEIR model in the Indian healthcare context, along with relevant resources and discussions on the default parameter choices. A static geo-located visualization is presented at an assumed 50% rate of hospital bed occupancy and default transmission rates that lead to *R*_0_ = 2. Self-updating waterfall plots are then offered for user-specified choices of region (national/state-specific), bed occupancy rate and transmission rates for different infection stages. The home page of the app also contains links to the data sources.

**Figure 9:**
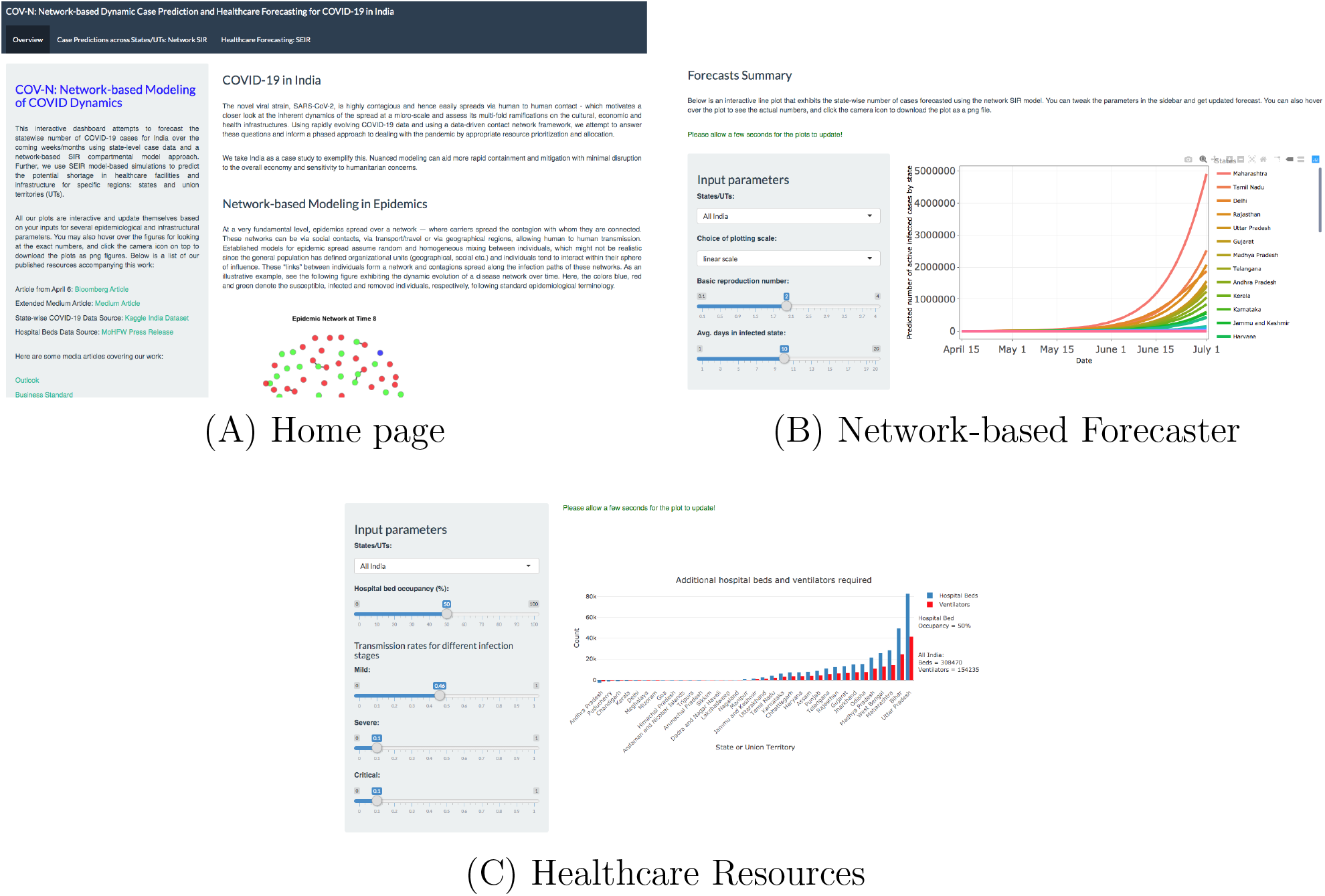
Representative snapshots of the R Shiny app providing interactive predictions and healthcare forecasts.

## 5 Discussion and conclusions

In this paper, we adapted a network-based SIR model to analyze regional contact networks and demonstrate the utility of such methods in the context of early stages of the COVID-19 spread in India. We believe such methods could be profitably used to inform policy-level decisions by allowing detection of hotspots and identification of key predictors characterizing these hotspots such as population density, co-morbidities, socio-economic and demographic factors, and access of health care facilities. Our hope is such nuanced modeling can aid more rapid and focused containment and subsequent mitigation with regional specificity, minimal disruption to the overall economy and sensitivity to humanitarian concerns.

The data and analyses in this article along with the interactive app can aid regional/state level specificity to allow a proportional response, prepare for the surge by planning for different scenarios, and create a road-map so the country can eventually come back to normalcy in a phased manner that is socially responsible, equitable, and sustained. Our research group provided some specific recommendations that were summarized in an opinion piece we wrote in April 2020.

There are several potential refinements and future directions possible with network-based SIR models. We used spatial/location information to construct our networks which, while admittedly limited, provided some broad clues regarding the early pandemic spread in India. The network reconstruction can be further improved by layering with more precise information that can be collected e.g. micro-level county/district level data, more efficient contact tracing channels, travel information etc. Given the limitations of our current dataset, we did not explore these avenues but these could provide valuable insights, prospectively or retrospectively, for learning the dynamics of the COVID-19 spread. In terms of methodology, there are several refinements possible. We only focused on simulation based on assumed parameter choices such as *R*_0_, and not on any formal inference or hypothesis testing. One interesting avenue could be to construct a unified framework, through likelihood based constructions for network-based models and employ a frequentist or Bayesian estimation procedure and obtain uncertainty quantifications. Furthermore, one could explore potential stochastic formulations of the deterministic SIR-type models that can accommodate flexible noise scenarios such as nonlinearity or non-stationarity (Bhadra et al., 2011).

When we started this work in March 2020, given the lack of tests and the large number of asymptomatic carriers, the strategy for slowing the spread of the COVID-19 pandemic had changed from containment to mitigation (Parodi and Liu, 2020). In essence, the focus was on slowing the further spread of the virus, reducing the anticipated surge in health care use, providing patients with the right level of care to maximize the likelihood that the majority of patients will only require time-limited home isolation, expanding testing capability to increase available hospital capacity, and tailoring isolation to minimize transmission of SARS-CoV-2. In a country like India density is ubiquitous given the population. The social fabric consists of extensive family and societal networks. The challenge is coming up with immediate responses, such as rapid lock-downs, that will have a lasting effect on many classes of society, especially the urban poor. What is needed is a resilient and sustainable approach, with regional sensitivity and national connectivity. Moving forward, a strategy is needed which makes India’s density its advantage in dealing with COVID-19 and future pandemics. Using technological infrastructures, such as leveraging the massive mobile network of India — ensures that the reminders about social distancing, hand-washing and hygiene are not overlooked. This can also ensure that testing is strategic (not possible to test everyone), contact tracing is immediate (once a person is tested positive it is relatively easy to trace previous contacts and isolate them immediately), and quarantine is smart (right level of quarantine based on the level of risk). In addition, one has to ensure that healthcare facilities are ready for the surge by decanting non-critical services, rapidly converting existing facilities, ramping up medical equipment and supplies, and leveraging alternate care facilities. The biggest challenge will be to do this in a humane, culturally sensitive and respectful way. We believe the strongest response to contagions is through building resilient communities that can manage the contact networks - and our models can potential aid this endeavor for current and future pandemics.

### Media coverage

Our work was highlighted in both national^15^ and local media^16^, especially in the context of the state of Madhya Pradesh and Indore district, where the second co-first author is based. The initial projections, along with the estimated pressure on the healthcare system helped the local administration to come up with mitigation strategies to contain the infection spread, simultaneously educating the common mass about the disease, and taking individual responsibilities from their end, like adoption of effective hygiene practices, wearing masks, avoiding social gatherings, among others.

## Data Availability

Individual-level, location-informed COVID-19 data for Indian states and union territories were obtained from the Kaggle database available at \url{https://www.kaggle.com/sudalairajkumar/covid19-in-india}. Data on state-wide daily infections is available from \url{https://api.covid19india.org/}.

https://www.kaggle.com/sudalairajkumar/covid19-in-india

https://api.covid19india.org/

https://bayesrx.shinyapps.io/COV-N/

## Declarations

## Availability of data and material

Individual-level, location-informed COVID-19 data for Indian states and union territories were obtained from the Kaggle database available at https://www.kaggle.com/sudalairajkumar/covid19-in-india. Data on state-wide daily infections is available from https://api.covid19india.org/.

## Authors’ contributions

S.B. and V.B. conceived the project. R.B., S.B., and V.B. developed the statistical framework and R.B. developed the software and performed statistical analysis. S.M. analyzed the healthcare forecasting data. All authors wrote, read and approved the final manuscript.

## Acknowledgements

We would like to thank Dr. Upali Nanda for her input in the initial stages of this work.

## Footnotes

1 https://knowindia.gov.in/profile/population.php

2 https://knowindia.gov.in/states-uts/

3 https://pib.gov.in/PressReleaseIframePage.aspx?PRID=1601095

4 https://www.covid19india.org/

5 https://www.worldometers.info/coronavirus/

6 https://www.who.int/news-room/commentaries/detail/transmission-of-sars-cov-2-implications-for-infection-prevention-precautions

7 https://www.kaggle.com/sudalairajkumar/covid19-in-india

8 https://pib.gov.in/PressReleasePage.aspx?PRID=1539877

9 Hindustan Times newspaper report, October 14, 2020, available at http://bit.ly/2Jl5qGG

10 The Times of India newspaper report, October 10, 2020, available at http://bit.ly/38AUKvY

11 https://www.bbc.com/news/world-asia-india-51997488

12 https://pib.gov.in/PressReleasePage.aspx?PRID=1539877

13 https://data.gov.in/resources/state-wise-population-decadal-population-growth-rate-and-population-density-2011-0

14 https://alhill.shinyapps.io/COVID19seir/

15 http://bit.ly/bus-std-report

16 http://bit.ly/free-press-report

